# Female Infertility and Neurodevelopmental Disorders in Children: associations and evidence for familial confounding in Denmark

**DOI:** 10.1101/2024.09.17.24313638

**Authors:** Khaoula Ben Messaoud, Vahe Khachadourian, Elias Arildskov, Stefan N. Hansen, Renee Gardner, Cecilia Ramlau-Hansen, Linda Kahn, Magdalena Janecka

## Abstract

**IMPORTANCE:** Existing research suggests the impact of infertility on the risk of neurodevelopmental disorders in children, however, studies to date have failed to separate the impact of male and female infertility, often blurring the lines with proxies that encompass all forms of infertility. Moreover, while both health conditions co-occurring with infertility and genetic factors operating upstream have been suggested to influence the association between infertility and child outcomes, their roles and potential impact on observed associations remain unclear.

**OBJECTIVE:** The objectives of this study are to investigate the relationship between female infertility and autism in the child, differentiating it from the effects of male and the couple infertility; consider the role of various maternal and birth factors in the association; and examine the effects of shared familial confounders on the association.

**DESIGN SETTING AND PARTICIPANTS:** Danish population-based cohort study, including all singleton live births in Denmark 1998-2015, their parents and parents’ siblings. The cohort was followed up until December 31, 2016.

**EXPOSURES:** The exposure was a history of female infertility in the mother and the mother’s sister. We examined four definitions of female infertility based on the ICD-10 codes derived from the Danish National Patient Register - any female infertility; specified female infertility; female exclusive infertility; and female or male infertility.

**MAIN OUTCOME AND MEASURES:** The outcome was diagnosis of autism spectrum disorder (ASD) in the Danish Psychiatric Central Research Register or the national patient register. A multivariable Cox regression model was used to estimate the associations between female infertility and autism, accounting for child’s sex, year of birth, maternal age, education level, chronic comorbidities, and pregnancy and birth complications. The effects of shared familial factors on the association were analyzed using exposure information from the child’s maternal aunt.

**RESULTS:** The cohort included 1,131,899 mother-child pairs, among which 18,374 children with ASD diagnosis. History of female infertility in the mother (all definitions) was significantly associated with autism in the child, with the association remaining robust after adjustment for covariates (HR_adj_=1.14 (95% CI, 1.03-1.26) for specified infertility). The diagnosis of infertility in a child’s maternal aunt was also significantly linked to the child’s autism risk, even after adjustment for maternal infertility (HR_adj_=1.10 (95% CI, 1.00-1.20).

**CONCLUSIONS AND RELEVANCE:** in This population-based birth cohort study, we found a slightly higher risk of autism in children born to mothers with a history of infertility, with the association remaining consistent across various definitions of female infertility and robust to adjustments for demographic, child, and maternal factors. The study suggests for the first time that shared familial factors, possibly both genetic and non-genetic, could be influencing both female infertility and the risk of autism in children, indicating a need for further investigation into these familial effects.

## Introduction

The prevalence of neurodevelopmental conditions in children has increased in recent years, reaching nearly 17% in the US between 2018 and 2021. Autism, usually manifesting in early childhood, is one of the most common neurodevelopmental condition, with a prevalence of ∼3% in children and adolescents in the US.^1^ In addition to this high, and increasing^2,3^ prevalence, recent studies have indicated that both autism and other neurodevelopmental disorders are associated with a large range of physical health problems across the lifespan.^4–7^

Involvement of genetic factors in autism etiology is well established.^8^ However, multiple non-genetic factors have also been associated with a higher likelihood of autism in the child, including perinatal complications such as pregnancy complications, preterm birth, and cesarean delivery.^9–12^ Additionally, maternal psychiatric, metabolic, and reproductive conditions, including depression^13^, obesity,^12,14^ and polycystic ovarian syndrome (PCOS),^15,16^ have also been associated with autism in offspring.^14,17,18^ However, the mechanisms underlying these associations, and the potential role of non-genetic factors in autism etiology remain unknown.

Many of these putative non-genetic factors are associated with an infertility diagnosis – either as a co-occurring condition, lifestyle risk factor, or consequence. Several studies have suggested that autism is more frequent in children born following an infertility treatment.^15,19,20^ However, other studies have indicated that these effects arise due to infertility, subfertility or anovulatory infertility, rather than the infertility treatment itself.^16,21,22^ For example, a Canadian population-based study by Velez et al. demonstrated a higher likelihood of autism following maternal infertility treatment (ovulation induction/ intrauterine insemination, in vitro fertilization (IVF)/intracytoplasmic sperm injection (ICSI)); however, children born after pregnancy with a history of infertility were equally likely to be diagnosed with autism, whether infertility treatment was used or not — suggesting a link between infertility and autism, independently of infertility treatment.^22^

Despite the robustness of Velez et al. results, infertility is a complex clinical phenomenon, leaving many questions about the association between parental history of infertility and risk of neurodevelopmental disorders in offspring. First, large, population-based studies to date have investigated the association between parental infertility and offspring outcomes without drawing a distinction between female- and male-factor infertility. Instead, to define infertility, these studies used seeking medical help for infertility problems or the time to pregnancy as proxies of *couple’s* infertility. As both of these measures capture simultaneously male, female and unexplained infertility,^21,22^ their use renders the attribution of the observed association to biological factors underlying either male or female infertility difficult. Next, the contribution of other health factors to the association between infertility and offspring neurodevelopment remains unclear. For example, maternal reproductive and metabolic conditions, as well as pregnancy complications are associated with both female infertility^23^ and neurodevelopmental outcomes in offspring. While they have previously been suggested to be mediators of the relationship between infertility and autism,^22^ the potential confounding effects of the medical conditions preceding the pregnancy cannot be excluded.^24^ Finally, the role of familial factors – whereby, for example, shared genetic liability for infertility and neurodevelopmental conditions could underlie the observational association between female infertility and autism in offspring – remains unexplored.

To assess the association between female infertility and autism, distinguish it from any association attributable to male infertility, and interrogate potential familial confounding, we designed a nationwide birth cohort study, including all live births between 1998 and 2015 in Denmark. To interrogate this association further, we considered a range of other maternal, pregnancy, and birth factors; differentiated between effects of different infertility types; and explored the risk of autism associated with infertility in second-degree relatives.

## Methods

### Study cohort

The cohort included all children born alive in Denmark between January 1^st^ 1998 and December 31, 2015, their parents, and their parents’ siblings identified in the Danish Medical Birth Registry (DMBR) and the Danish Central Population Register (DCPR).^25–27^ All individuals were followed up until December 31, 2016. Exclusion criterion was maternal age <13 or >55 years. The study followed the Strengthening the Reporting of Observational Studies in Epidemiology (STROBE) reporting guidelines.^28^

### Exposure

The exposure was defined as maternal history of infertility. Four non-mutually exclusive categories of female infertility were defined based on the information about maternal and paternal infertility status derived from the diagnostic data available in the Danish National Patient Register (DNPR) within three years before the child’s birth^27^:*Any female infertility, specified female infertility, female exclusive infertility, female or male infertility* (**Table 1**). All diagnoses were classified based on the *International Statistical Classification of Diseases and Related Health Problems, Tenth Revision (ICD-10)*, which has been in place in Denmark since 1994. Infertility diagnosis in mother’s sister was defined using the same diagnostic codes but ascertained between four years prior to the birth of the index child and the end of follow-up, to capture the familial, time-invariant signal.

**Table 1:** Infertility exposure definition.

| Infertility definition | ICD-10 codes |
| --- | --- |
| *Any female infertility* | At least one of the following codes: N97.0, N97.1, N97.2, N97.3, N97.8, N97.9 |
| *Specified female infertility* | At least one of the following codes: N97.0, N97.1, N97.2, N97.3, N97.8 |
| *Female exclusive infertility* | At least one of the following codes: N97.0, N97.1, N97.2, N97.3, N97.8, or N97.9<br>With non-occurrence of: N97.4, N46.0, N46.1, N46.8, N46.9 |
| *Female or male infertility* | at least one of the following codes in the mother: N97.0, N97.1, N97.2, N97.3, N97.8, N97.9 or N46.0, N46.1, N46.8, N46. |

### Outcomes

We ascertained autism cases using diagnosis of autism spectrum disorder (ASD) in the child during the follow up period, from birth until December 31, 2016. The relevant ICD-10 diagnosis codes recorded in the Danish Psychiatric Central Research Register (DPCRR)^26^ or DNPR^27^ were used to identify autism cases: F84.0, F84.1, F84.5, F84.8, or F84.9, excluding F84.2 and F84.3. Diagnoses from inpatient, outpatient, and emergency room visits have been reported to the DPCRR since 1995. The validity of the ASD diagnosis in Danish registries has been verified.^29^

### Covariates

We considered a range of potential confounders and precision variables in the association between maternal history of infertility and child autism, including family socio-demographic variables, maternal chronic comorbidities, and pregnancy and birth complications. To focus on the most common maternal health conditions, ICD-10 codes with fewer than 10 occurrences in the cohort were excluded.

#### Socio demographic covariates

In all models we adjusted for child’s sex, year of birth and *maternal age* at child’s birth, which is associated with both female infertility^30^ and autism in the child^31^. Maternal age was entered into the model as restricted cubic spline with 4 knots. *Maternal education* was then included as an indicator of family’s socio-economic status, which is also associated with both maternal and child diagnoses; maternal education was classified into 4 levels: primary and lower secondary (reference), upper secondary to secondary vocational education, short-cycle tertiary education to bachelor level, and long tertiary education (master’s to PhD level).

#### Chronic comorbidities

We accounted for a range of maternal comorbidities associated with both female infertility and the risk of autism, including: reproductive and metabolic disorders (*PCOS (*E28.2*), obesity (*E65-E68*), diabetes (*E08-E13*), thyroid disorders (*E01; E02; E04-E07), and *other metabolic disorders (*E70-E88)) present during the three years prior to the child’s birth; and maternal mental health disorders (*schizophrenia (*F20, F21), *mood disorders (*F31-F34), *anxiety (*F41-F43*), alcohol-related disorders (*F10*), eating disorders (*F50*), specific personality disorders (*F60*), and mental disorder not specified (*F99)) diagnosed during the pregnancy or in the three years leading up to the birth. All maternal diagnoses were identified using diagnostic data available in the DNPR and DPCRR.

#### Pregnancy complications

Pregnancy complications were identified in the 12 months before child’s birth and included: *Common Pregnancy complications* (*multiple gestation* (O30), *maternal care for known or suspected fetal abnormality and damage* (O35), *maternal care for other fetal problems* (O36), *premature rupture of membranes* (O42), *false labor* (O47)); *other maternal diseases complicating pregnancy* (O99); *other abnormal products of conception* (O02); *preeclampsia (*O14); *Other maternal disorders related to pregnancy* (abnormal findings on antenatal screening of mother (O28),and diabetes mellitus in pregnancy (O24)).

#### Birth complications

We adjusted for *Cesarean delivery* (O82), *labor complications* (abnormalities of forces of labor (O62), labor and delivery complicated by abnormality of fetal acid-base balance (O68), postpartum hemorrhage (O72)).

### Statistical analysis

We performed a multivariable Cox regression model to estimate hazard ratios (HRs) and 95% confidence interval (CIs) for the associations between maternal infertility and autism, using the child’s attained age in days as the time scale. All children were followed up through 2016, until first autism diagnosis, death, or emigration out of Denmark, whichever occurred first. To account for within-family correlations due to the presence of siblings in the dataset we estimated robust standard errors. The models were stratified by the child’s year of birth and adjusted for potential confounders, employing a stepwise adjustment approach. Initially, we adjusted for the child’s sex and the maternal age. Subsequently, we included adjustments for the maternal education level, and then for additional factors including chronic comorbidities, as well as pregnancy and birth complications. We ran the model for the four definitions of the exposure (“any female infertility”, “specified female infertility”, “female exclusive infertility” and “female or male infertility”, as defined in Table 1).

To assess the effects of shared familial confounders on the association between infertility and autism in the child, we estimated autism risk associated with a diagnosis of specified female infertility in child’s maternal aunt (mother’s sister). We restricted the analytical sample to cohort children whose mothers had at least one sister in the source population. To estimate the association between the aunt’s specified infertility status and autism likelihood in the child, we followed the same stepwise adjustment sequence as for maternal exposure. Standard errors were calculated using bootstrapping, by computing a coefficient for each one of the 1000 bootstrap samples. Due to potential familial co-occurrence of infertility among family members (child’s mother and aunt), we conducted additional analyses controlling for specified maternal infertility status.

Analysis was restricted to individuals with complete covariate data. All analyses were performed using R Statistical Software (v4.3.2; R Core Team 2023).

## Results

### Descriptive analysis

There were 1,131,899 mother-child pairs in the cohort, including 18,374 (1.62%) children diagnosed with ASD before the end of the follow-up period. **Error! Reference source not found**.**Table 2** describes the cohort by child’s autism diagnosis status.

**Table 2:** Characteristics of 1 131 899 live births in Denmark, 1998 to 2015, according to the occurrence of ASD diagnosis in the child.

| Characteristics | ASD diagnosed<br>(n=18374) | Non-ASD diagnosed<br>(n=1113525) | Total<br>(n=1131899) |
| --- | --- | --- | --- |
| **Maternal age at birth, mean (SD)** | 30.3 (5.1) | 30.6 (4.9) | 30.6 (4.9) |
| **Maternal educational status n (%)** |  |  |  |
| Primary & lower secondary | 4828 (26.3) | 211468 (19.0) | 216296 (19.1) |
| Upper to vocational secondary education | 7844 (42.7) | 447248 (40.2) | 455092 (40.2) |
| Short-cycle tertiary to bachelor | 3865 (21.0) | 292433 (26.3) | 296298 (26.2) |
| Long tertiary education (masters) to PhD | 1217 (6.6) | 106567 (9.6) | 107784 (9.5) |
| Missing | 620 (3.4) | 55809 (5.0) | 56429 (5.0) |
| **Mother with at least two children in the cohort n (%)** |  |  |  |
| No | 5229 (28.5) | 275100 (24.7) | 280329 (24.8) |
| Yes | 13145 (71.5) | 838425 (75.3) | 851570 (75.2) |
| **Child sex** |  |  |  |
| Female | 4509 (24.5) | 546916 (49.1) | 551425 (48.7) |
| Male | 13865 (75.5) | 566609 (50.9) | 580474 (51.3) |
| **Child year of birth n (%)** |  |  |  |
| 1998 | 1750 (9.5) | 64381 (5.8) | 66131 (5.8) |
| 1999 | 1875 (10.2) | 64365 (5.8) | 66240 (5.9) |
| 2000 | 1936 (10.5) | 65175 (5.9) | 67111 (5.9) |
| 2001 | 1754 (9.5) | 63695 (5.7) | 65449 (5.8) |
| 2002 | 1631 (8.9) | 62484 (5.6) | 64115 (5.7) |
| 2003 | 1526 (8.3) | 63133 (5.7) | 64659 (5.7) |
| 2004 | 1333 (7.3) | 63358 (5.7) | 64691 (5.7) |
| 2005 | 1237 (6.7) | 63123 (5.7) | 64360 (5.7) |
| 2006 | 1147 (6.2) | 63891 (5.7) | 65038 (5.7) |
| 2007 | 1010 (5.5) | 63200 (5.7) | 64210 (5.7) |
| 2008 | 838 (4.6) | 64248 (5.8) | 65086 (5.8) |
| 2009 | 732 (4.0) | 62203 (5.6) | 62935 (5.6) |
| 2010 | 648 (3.5) | 62879 (5.6) | 63527 (5.6) |
| 2011 | 460 (2.5) | 58662 (5.3) | 59122 (5.2) |
| 2012 | 289 (1.6) | 57762 (5.2) | 58051 (5.1) |
| 2013 | 142 (0.8) | 55814 (5.0) | 55956 (4.9) |
| 2014 | 53 (0.3) | 56874 (5.1) | 56927 (5.0) |
| 2015 | 13 (0.1) | 58278 (5.2) | 58291 (5.1) |

### Association between female infertility and offspring autism

Maternal history of infertility was significantly associated with autism in the child across the four different definitions of exposure (specified female infertility (HR_adj_=1.20, 95% CI: 1.09-1.32), any female infertility (HR_adj_=1.10, 95% CI:1.02-1.17), female exclusive infertility (HR_adj_=1.11, 95% CI:1.02-1.20); and female or male infertility (HR_adj_=1.16, 95% CI:1.09-1.23)) after adjustment for child sex, maternal age, and maternal educational status. The association estimates were not substantially different across the exposure definitions (Figure 1).

**Figure 1:**
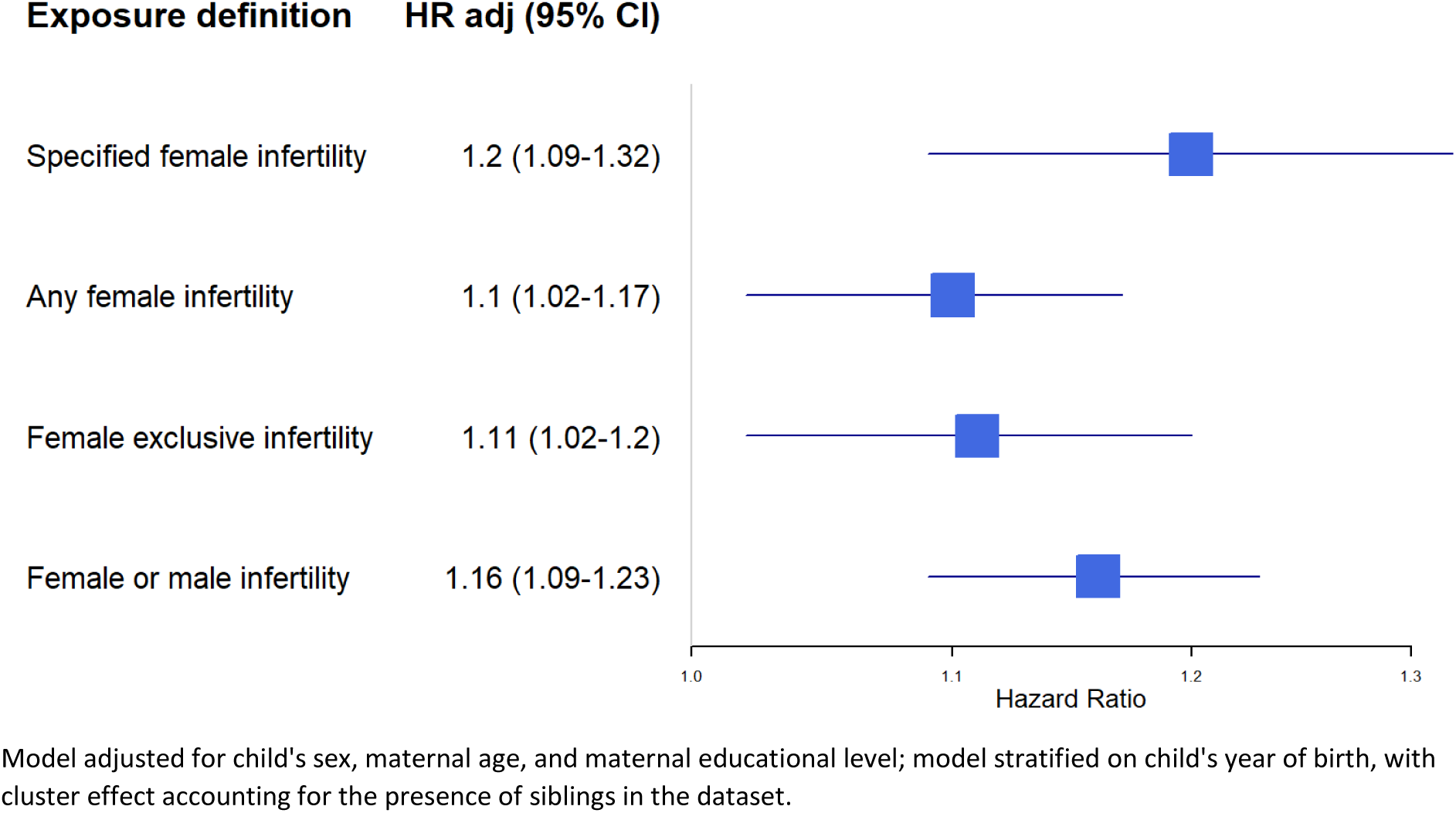
The adjusted risk of ASD in the child according to the female infertility exposure.

The associations between female infertility and offspring autism were robust to adjustment for potential confounders. For specified female infertility, results from the model adjusted for maternal age and child sex (HR=1.19, 95% CI: 1.08-1.31) remained consistent through additional adjustment for maternal education (HR=1.20 (95% CI: 1.09-1.33)) and chronic comorbidities, pregnancy- and birth-complications (HR=1.14 (95% CI: 1.03-1.26)). Results from the fully adjusted model are presented in Table 3.

**Table 3:**
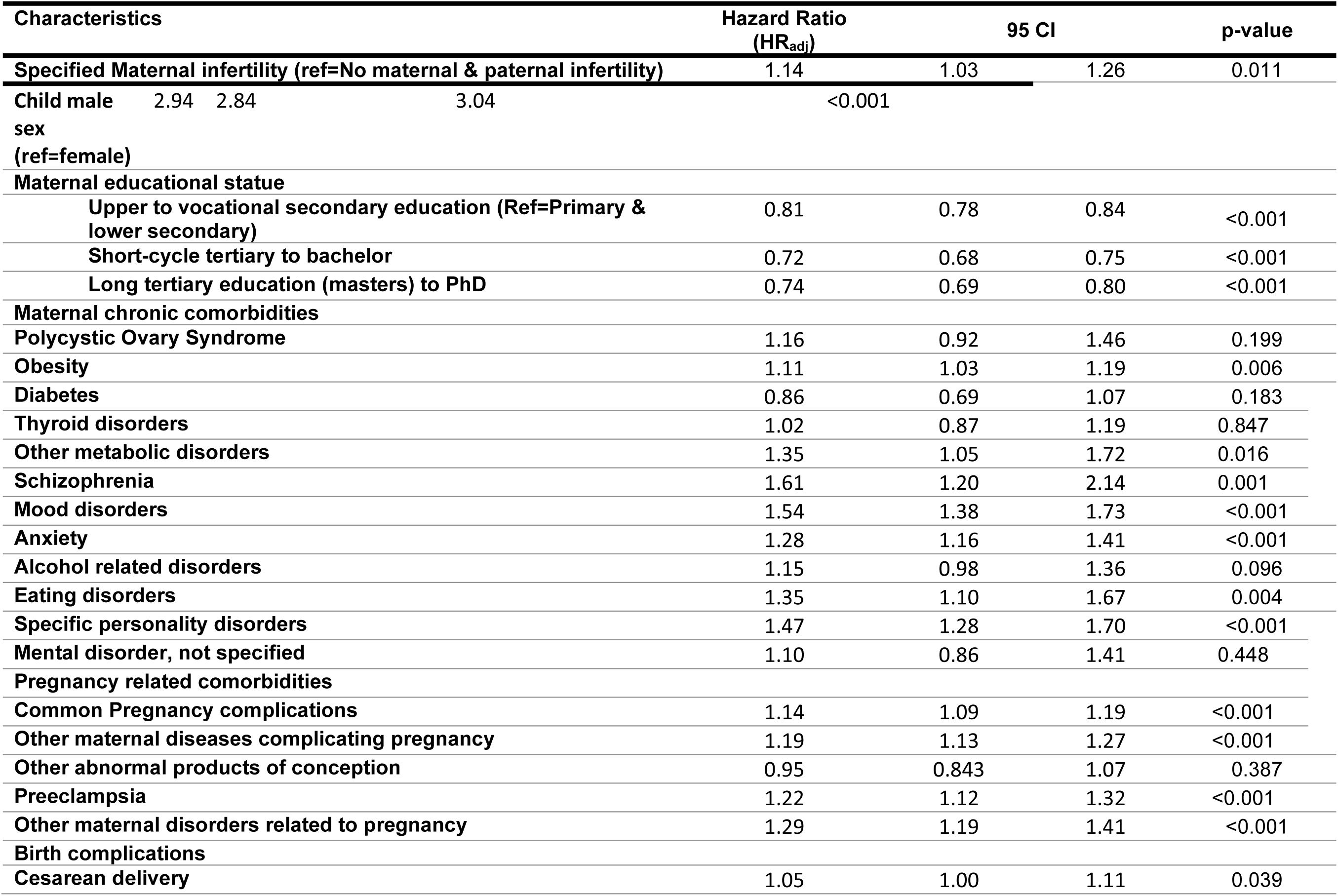
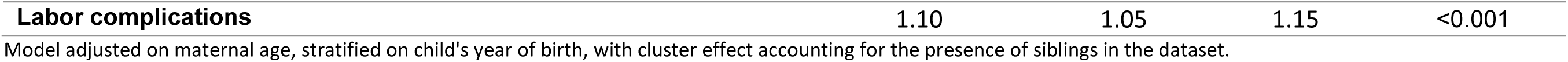
Risk of Autism Spectrum Disorder (ASD) by Maternal, Pregnancy-Related, Obstetric, and Child-Related Factors.

### Familial effects in the association between infertility and autism

Maternal aunt’s diagnosis of specified female infertility was significantly associated with autism in the child, after adjustment for child’s sex, maternal age and educational status (HR=1.10, 95% CI: 1.00-1.20). These effects remained significant, and unchanged after additional adjustment for specified maternal infertility (HR=1.10, 95% CI: 1.00-1.20).

## Discussion

### Main findings

In this population-based birth cohort study, we found a slightly higher risk of autism in children of mothers with a history of infertility. This association was observed across all explored definitions of female infertility (any female infertility, specified female infertility, female exclusive infertility, female or male infertility), and was robust to adjustment for a range of demographic, child, and maternal covariates. Leveraging the extensive family linkage in the Danish register, we were able to demonstrate that this association arises at least in part due to familial factors.

While we observed that maternal conditions co-occurring with infertility, including obesity, metabolic conditions, and pregnancy complications, were themselves significantly associated with autism, the associations between maternal history of infertility and autism observed in our sample were not driven by those effects. Although adjustment for these factors resulted in a subtle attenuation of the main effect estimate, the association remained statistically significant. Nevertheless, while this adjustment did not affect our conclusions, future studies should investigate whether maternal pregnancy and birth complications should be included as covariates given their potential mediating role in the association between maternal infertility and offspring development.

Importantly, both maternal and maternal aunt’s diagnosis of specified female infertility was associated with autism in our sample. As the aunt’s diagnosis has no direct impact on the index child, these results suggest that the original association arose, at least in part, due to the familial factors associated with infertility shared between the mother and her sister. These familial effects could include both genetic and non-genetic factors that tend to be shared within broader family pedigrees (e.g., area-level pollution). The magnitude of the association between infertility and autism was 50% lower for aunt’s vs maternal infertility — consistent with the 50% lower relatedness between the child and their aunt vs mother, and thus providing support for the genetic origin of these effects. Nevertheless, the exact sources of the familial effects observed in our study remain to be established.

### Comparison with prior studies

A Canadian population-based study also reported a higher risk of autism in children born to couples with a history of infertility, a risk that was partially mediated by certain pregnancy, obstetrical, and perinatal factors.^22^ In contrast to our study, Velez et al. did not specifically investigate infertility of female origin. Nevertheless, their estimate of the association between offspring ASD in child and couple history of infertility. study was similar to ours (HR_adj_=1.20, 95% CI: 1.15-1.25) vs (HR_adj_=1.15, 95% CI: 1.03-1.26).

Considering the impact of parental history of infertility on the risk of autism in the child, previous studies primarily focused on effects of the use of assisted reproductive technologies. While these studies reported an increased risk of autism in children among treated couples,^33^ the estimates derived in these studies did not differentiate between the role of treatment and the infertility itself. However, Velez et al. demonstrated that the effects of infertility on neurodevelopment are more likely attributable to the infertility diagnosis, rather than treatment, as the adjusted risk of autism did not differ between the couples who were subfertile but non-treated (HR=1.20 (95% CI, 1.15-1.25)) and those treated with ovulation induction/ intrauterine insemination (HR=1.21 (95% CI, 1.09-1.34)) or IVF/ ICSI (HR=1.16 (95% CI, 1.04-1.28)).^22^ A common functional pathway between autism and female infertility could involve folate and/or hormonal signaling, both of which have a demonstrated impact on female fertility^34^ and neurodevelopment,^35^ however, the exact mechanisms remain to be elucidated.

Additional adjustment for pregnancy and birth complications resulted in lower point estimates, however, the results remained statistically significant.^36,16^ As these perinatal and birth complications have been identified as potential mediators in the association between maternal infertility and autism, it remains to be investigated whether adjustment for these factors in our study represents a correct analytical decision. However, this adjustment did not impact our conclusions, and our analyses show that even if such mediation occurs, it is incomplete, and the association between female infertility and autism remains significant even after adjustment for these pregnancy and birth factors.

### Strengths and limitations

The strengths of this study include a large, population-based sample, robust family linkage, long follow-up, and adjustment for a broad spectrum of covariates. To the best of our knowledge, this study is first to comprehensively explore the association between female – as opposed to couple’s – infertility with child’s autism and consider a range of definitions for maternal infertility. The robustness of our results is underscored by the strength and consistency of our findings, regardless of the proxy used for female infertility exposure definition. Finally, multi-generation family linkage in the Danish register has enabled us to identify parental siblings and conduct the analyses beyond nuclear families, providing novel evidence for the role of familial factors in the association between female infertility and autism.

However, the study also has certain limitations, including lack of consideration of infertility treatment. Although the study by Velez et al., based on 1.3 million children, showed that the underlying infertility itself, rather than treatment, is associated autism,^22^ we did not verify these findings in our data. We also excluded unexplained infertility in the couples, which may include some cases of unidentified female infertility. Furthermore, our cohort may include a residual number of females who received eggs from a donor, which could have affected the familial comparisons.

## Conclusions

This study advances our understanding of the association between female infertility and autism in children. Our findings indicate a slightly higher risk of autism in children born to mothers with a history of female infertility, a pattern that is consistent across various definitions of female infertility. This study also suggests, for the first time, that shared familial factors could be influencing both female infertility and the risk of ASD in children, warranting their further investigation.

## Ethics statement

Under the Danish legislation, the national register data are available for research without ethical approval or informed consent from individuals included in the database. To ensure compliance with the Danish law and individual privacy, each project is approved by the Danish Health Data Authority.

## Data availability

The Danish Scientific Ethical Committee system, as well as all relevant register authorities, including the Danish Data Protection Agency, Statistics Denmark, and the Danish Health Data Authority, approved access to these data under strict conditions regarding access and data export. Under these conditions there are no provisions for exporting individual level data, all or in part, to another institution, in or outside of Denmark. All summary statistics of the measures of associations used to draw the study conclusions are presented in the supplemental material. The corresponding author will respond to any potential additional queries.

## Author contributions

KB, VK and MJ designed the study; KB, VK and MJ planned the analysis. EA carried out the analysis. KB prepared the first draft of the manuscript. MJ obtained the funding and provided supervision. All authors contributed to interpretation of the findings, and critical revisions.

